# Forecasting the Worldwide Spread of COVID-19 based on Logistic Model and SEIR Model

**DOI:** 10.1101/2020.03.26.20044289

**Authors:** Xiang Zhou, Xudong Ma, Na Hong, Longxiang Su, Yingying Ma, Jie He, Huizhen Jiang, Chun Liu, Guangliang Shan, Weiguo Zhu, Shuyang Zhang, Yun Long

## Abstract

**Background:** With the outbreak of coronavirus disease 2019 (COVID-19), a sudden case increase in late February 2020 led to deep concern globally. Italy, South Korea, Iran, France, Germany, Spain, the US and Japan are probably the countries with the most severe outbreaks. Collecting epidemiological data and predicting epidemic trends are important for the development and measurement of public intervention strategies. Epidemic prediction results yielded by different mathematical models are inconsistent; therefore, we sought to compare different models and their prediction results to generate objective conclusions.

**Methods:** We used the number of cases reported from January 23 to March 20, 2020, to estimate the possible spread size and peak time of COVID-19, especially in 8 high-risk countries. The logistic growth model, basic SEIR model and adjusted SEIR model were adopted for prediction. Given that different model inputs may infer different model outputs, we implemented three model predictions with three scenarios of epidemic development.

**Results:** When comparing all 8 countries’ short-term prediction results and peak predictions, the differences among the models were relatively large. The logistic growth model estimated a smaller epidemic size than the basic SERI model did; however, once we added parameters that considered the effects of public health interventions and control measures, the adjusted SERI model results demonstrated a considerably rapid deceleration of epidemic development. Our results demonstrated that contact rate, quarantine scale, and the initial quarantine time and length are important factors in controlling epidemic size and length.

**Conclusions:** We demonstrated a comparative assessment of the predictions of the COVID-19 outbreak in eight high-risk countries using multiple methods. By forecasting epidemic size and peak time as well as simulating the effects of public health interventions, the intent of this paper is to help clarify the transmission dynamics of COVID-19 and recommend operation suggestions to slow down the epidemic. It is suggested that the quick detection of cases, sufficient implementation of quarantine and public self-protection behaviors are critical to slow down the epidemic.

## Introduction

A novel coronavirus caused pneumonia cases in Wuhan, a city in Hubei Province in China,, beginning at the end of 2019. Governments had implemented various measures to protect their cities or countries, such as traffic restrictions, quarantine requirements for travelers, and contact tracing, However, a large-scale global movement of the population still inevitably caused the rapid spread of the disease, resulting in an epidemic throughout China and worldwide. In February 2020, the World Health Organization named the disease COVID-19, which stands for coronavirus disease 2019^1^. The virus that causes COVID-19 is named severe acute respiratory syndrome coronavirus 2 (SARS-CoV-2). The SARS-CoV-2 virus spreads mainly between people who are in close contact with one another and through respiratory droplets produced when an infected person coughs or sneezes^2^. The global spread of COVID-19 caused a surge in Asia, Europe, the Middle East and North America. As of March 11, 2020, with the global risk continuously increasing, there were already more than 118,000 cases in 114 countries, and 4,291 people have lost their lives. The WHO has characterized COVID-19 as a pandemic^3^.

With the number of cases growing in more than 150 countries and regions, modeling the transmission dynamics and estimating the development of COVID-19 are crucial to providing decisional support for public health departments and healthcare policy makers. Mathematical models are widely used in evaluating epidemic transmissions, forecasting the trend of disease spread, and providing optimal intervention strategies and control measures. A considerable number of recent studies have contended to estimate the scale and severity of COVID-19, and several mathematical models and predicting approaches have attempted to estimate the transmission of COVID-19^4-8^. The majority of the studies have estimated the basic reproductive number R_0_, a key parameter to evaluate the potential for COVID-19 transmission. However, different models often yield different conclusions in terms of differences in model structure and input parameters. It is imperative and critical to improve the early predictive and warning capabilities of potential models for the pandemic.

In the face of this new infectious disease and its complicated features with many unknown factors, single model estimations may infer biased results; therefore, we tried to make overall rigorous estimations by comparing different model results. To achieve an objective estimation, we investigated and implemented the two most common approaches and one extended approach: the logistic model, the susceptible-exposed-infected-removed (SEIR) model and the adjusted SEIR model. Countries had different initial times and levels of interventions and measures to reduce the risk of domestic secondary infections of COVID-19. We compared the models that have taken these effects into account or not, predicted the spread of the epidemic, and tried to compare different recommendations from three different models.

## Methods

We collected data on the epidemic situation of COVID-19 in eight high-risk countries located on three continents and compared the results with those of the logistic model and the SEIR model with different parameter setting scenarios. Data are from the Coronavirus COVID-19 Global Cases published by the Center for Systems Science and Engineering (CSSE) of Johns Hopkins University^9^. We used the existing reported data from January 23 to March 20, 2020, for observing, performing parameter estimation, and forecasting COVID-19 dynamics in different countries/regions.

### Logistic growth model and parameter estimates

In Scenario 1, we assume that the epidemic trend obeys a logistic growth curve. We used a logistic model to predict the disease trends. The logistic model’s essence is that curve fitting and its prediction results highly depend on historical data. It has often been used in the prediction of epidemic dynamics in previous studies^4,10,11^. Mathematically, the logistic model describes the dynamic evolution of infected individuals being controlled by the growth rate and population capacity. According to the following ordinary differential equation (a), we will obtain the logistic function (b). The model describes the dynamic evolution of the reported number of confirmed cases P being controlled by the growth rate r, and the initial value of P_0_ is the confirmed number of cases when T=0. The maximum case volume in the environment is K, which is the limit that can be reached by increasing to the final value of P(t), and r is the growth rate. We used the least squares method to fit the logistic growth function and then to predict the number of future confirmed cases. Since the case numbers reported at very early stages are usually inaccurate or missing, the initial date of the model was set as the day since the 100th confirmed case was reached.

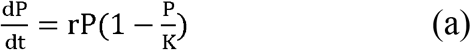

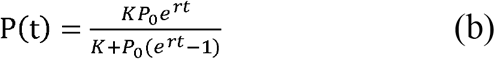

To evaluate the logistic model’s ability to predict infectious diseases such as COVID-19, we fit the logistic curve every 10 days since the day the 100th confirmed case was reached, and each time, we made 7-day predictions and used reported data for evaluation. In the experimental analysis, for each time of perdition, we obtained different result errors, as listed in Appendix A. As the number of confirmed cases increases, the predicted following 7 days of infections and future peak size and peak time are constantly changing. In addition, the shape of the curve will probably change due to exogenous effects, such as a new burst of infection, the implementation of control measures and public behaviors.

### SEIR model and parameter estimates

Based on the epidemiological characteristics of COVID-19 infection, the SEIR model is more commonly adopted to study the dynamics of this disease. SEIR is a deterministic metapopulation transmission model that simulates each individual in the population as a separate compartment, with the assumption that each individual in the same compartment has the same characteristics. By plugging in different settings of parameters, however, the models yield different results, and we compared their results to observe patterns of the COVID-19 spread under two different scenarios, namely, the basic SEIR (Scenario 2: without any interventions and measures) and the adjusted SEIR (Scenario 3: with strict interventions and measures).

In Scenario 2, we only used the basic SEIR model, and the population is divided into four classes: susceptible (S), exposed (E), infectious (I) and removed (R). The essence of the SEIR model is a system of ordinary differential equations over time. The disease trend it predicts only depends on parameters and the start time. The model is measured by the equation below, and the entire population was initially susceptible, with the assumption that all people have no immunity against COVID-19. The initial number of cases was collected from the reported data. Because reliable data are still scarce during the early days of a new outbreak, the initial date of the model was set as the day since the 100th confirmed case was reached for each country, which indicates different initial dates of the 8 observed countries.

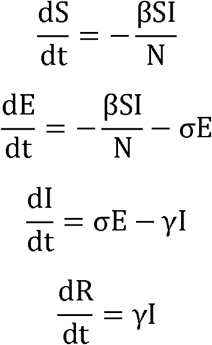

where S is the number of individuals in the susceptible population, E is the number of those in the exposed population, I is the number of those in the infected population, R is the number of recoveries or deaths, N is the number of those in the whole population, and β = k ∗ b is the product of the people exposed to the infected population k and the probability of transmission b. γ=1/D is the average rate of recovery or death in infected populations, where D is the average duration of the infection, and σ is the rate at which exposed individuals develop into those with infections.

In Scenario 3, we further used an adjusted SEIR model for COVID-19 estimation. According to recent studies, some investigations was conducted to measure the contribution to epidemic dynamics by public health intervention factors, such as the government locking down cities, taking measures to track and quarantine people who have close contact with confirmed cases, advocating citizens to maintain social distance and wash their hands frequently, a conclusion was yield that it will limit the number of confirmed cases 96% fewer than expected in the absence of interventions by implementing multiple measures together and interactively^12^. Therefore, this adjusted SEIR model considered the contact rate and quarantined proportion of COVID-19 transmission and divided the population into seven classes: susceptible, exposed, infectious, removed, quarantined susceptible, quarantined exposed and quarantined infected. A fraction of the susceptible population was quarantined and identified as Sq, and a fraction of the exposed population was isolated and identified as Eq. We provide their detailed equation as follows^8^:

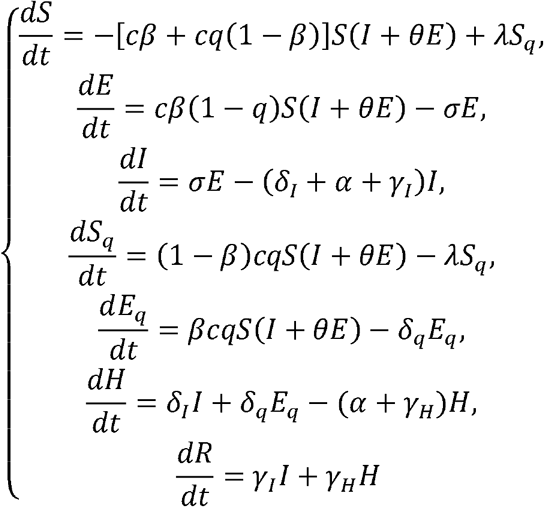

All countries reduced social contact among its people, although by implementing multiple different control measures, we give a preliminary average estimation for the contact rate and quarantined proportions as model parameters. In addition, as the quarantine and control measures take effect after a period of increasing cases, it has typically surged into a considerable number; therefore, the initial dates of 8 observed countries used in the adjusted model were different, and they were defined as the dates when the countries’ governments implemented strict interventions and control measures for a large-scale population. We assumed that the contact rate decreased since the government implemented these strict control measures. In terms of our previous study and related studies^8^, the contact rate was below 8 after a large-scale intervention was initiated. The initial populations of each country were acquired from published data^13^, and the initial infected and recovered population was set based on the reported data^9^. Model parameters are estimated on the basis of fitting reported data from the initial date, and the probability of transmission per contact of each country was estimated using early-stage data from each country based on Monte Carlo simulation. We assumed that the median incubation period was 5-6 days (ranging from 0-14 days) based on the WHO report^14^, the quarantine proportion of uninfected susceptible individuals was 60%∼80% of the population (under the assumption that a strict large-scale quarantine policy is executed) with the period set as 28 days, and the mortality rate was derived from reported data of each country. Based on the above assumptions, we implemented the adjusted SEIR model to obtain the minimal estimation.

### Spreading potential evaluation

To disclose the epidemic growth potential of each country, we used two parameters for the assessment. First, the greater the growth rate of the logistic curves is, the faster the curve grows. Second, the basic reproduction number R_0_^15^. We estimated the early transmission R_0_ of each country using the 10 days reported data since the 100th confirmed case was reached. We used the next-generation matrix to derive a formula for the R_0_^16^, as follows:

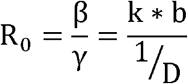

where β is the product of the population exposed to the infected population (k) and the probability of transmission (b). γ=1/D is the average rate of recovery or death in infected populations, where D is the average duration of the infection. In terms of related studies for the COVID-19 infection assumption^7^, we set D=7 to obtain the R_0_ estimation.

## Results

### COVID-19 Epidemic Progressing

We summarized the epidemic curves of the 8 observed countries to help clarify the global trends and disclose the spread pattern of different countries. Cases increased rapidly in these countries, including Italy, South Korea, Iran and France. The epidemic curves are shown in Figure 1. The initiation date of the curves was set as the earliest day since the 100th confirmed case was reached for 8 countries, which was February 20, 2020 as of South Korea (the initial dates for other countries can be found in Appendix B). However, as the epidemic curve follows the rule of rising, peaking, and then declining, during our observed period, all 8 countries are in their speedy rising stages but have not yet reached their peak and decline stages. Among these countries, the number of confirmed patients in Italy is the largest. After a small burst, as a result of a series of emergency prevention and control measures, the number of confirmed diagnoses in South Korea has increased slowly. Iran, Germany, Spain and the US have approximately 20,000 confirmed patients as of March 20, and there are no trends of a slowdown. Iran has more mild patients, so cure rates are higher. The confirmed cumulative number of patients in Japan is 963 as of March 20, and the number of cured patients is 191, which is the smallest number of cases in these 8 countries.

**Figure 1.**
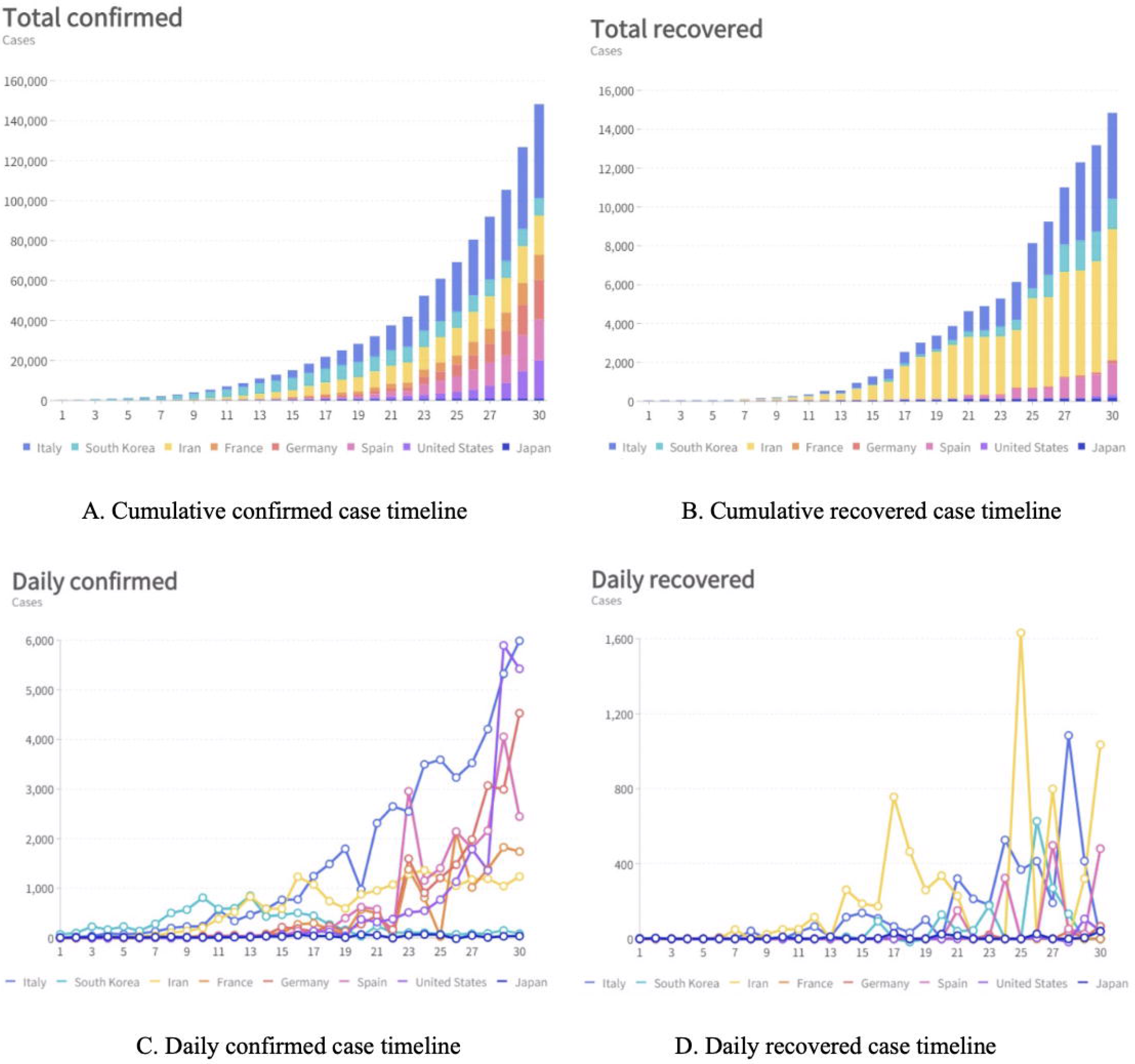
Epidemiological curves of the 8 observed countries(starting on February 20, 2020)

In Figure 2, the death tolls in Italy due to COVID-19 reached 4032 as of March 20, and the mortality rate of Italy reached 8.57%. Iran’s early negligence also caused a very high mortality rate of 40% at its early stage. Some countries also have high mortality rates, such as the US and Japan. However, South Korea and Germany controlled the mortality rate at approximately 1%, which is a positive indication.

**Figure 2.**
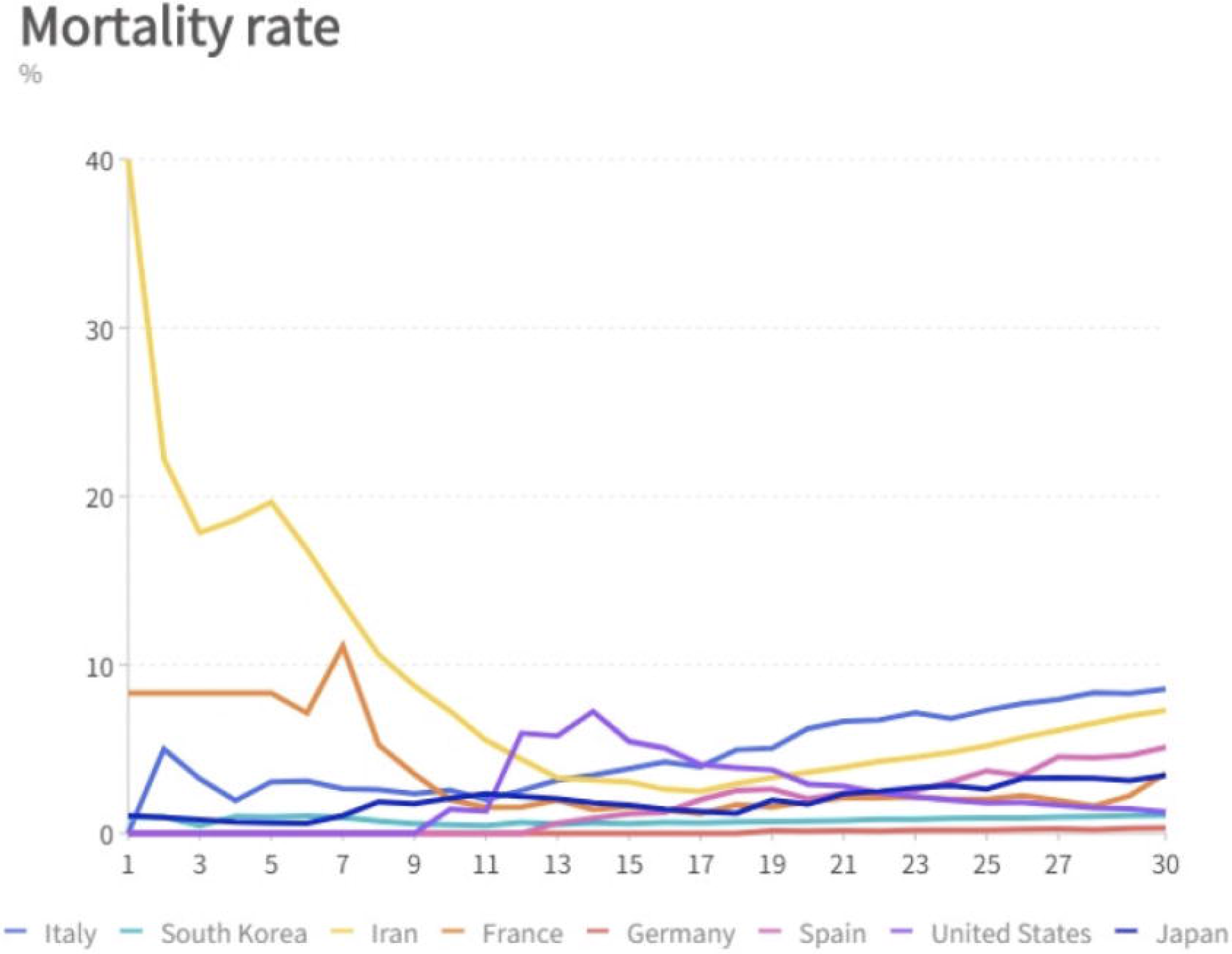
Mortality rate of the 8 observed countries (starting on February 20, 2020)

### Model Predictions

When contrasting the results from the three prediction models, we achieved quite different results for COVID-19 development, as shown in Table 1 and Table 2. Because different models were built on different theories and assumptions, their output measurements were varied, the cumulative number was used for the logistic model, and the active number was measured by SEIR models. The results disclosed the differences in the three mathematical models and further disclosed the prediction differences without/with the consideration of interventions.

**Table 1.** Short-term epidemic predictions of the 8 countries

| Countries<br>Models |  | Italy | Iran | South<br>Korea | Germany | France | US | Spain | Japan |
| --- | --- | --- | --- | --- | --- | --- | --- | --- | --- |
| Logistic<br>model<br>(Scenario 1) | 7-day<br>prediction<br>(cumulative) | 75,406 | 22,319 | 8139 | 32,594 | 24,250 | 272,880 | 26,692 | 1281 |
| Basic<br>SEIR<br>(Scenario 2) | 7-day<br>prediction<br>(active) | 22,620 | 27,840 | 42,480 | 51,820 | 71,830 | 79,120 | 12,040 | 1235 |
| Adjusted<br>SEIR<br>(Scenario 3) | 7-day<br>prediction<br>(active)(active) | 70,120 | 34,040 | 10,210 | 67,640 | 24,950 | 34,810 | 58,020 | 1540 |

**Table 2.**
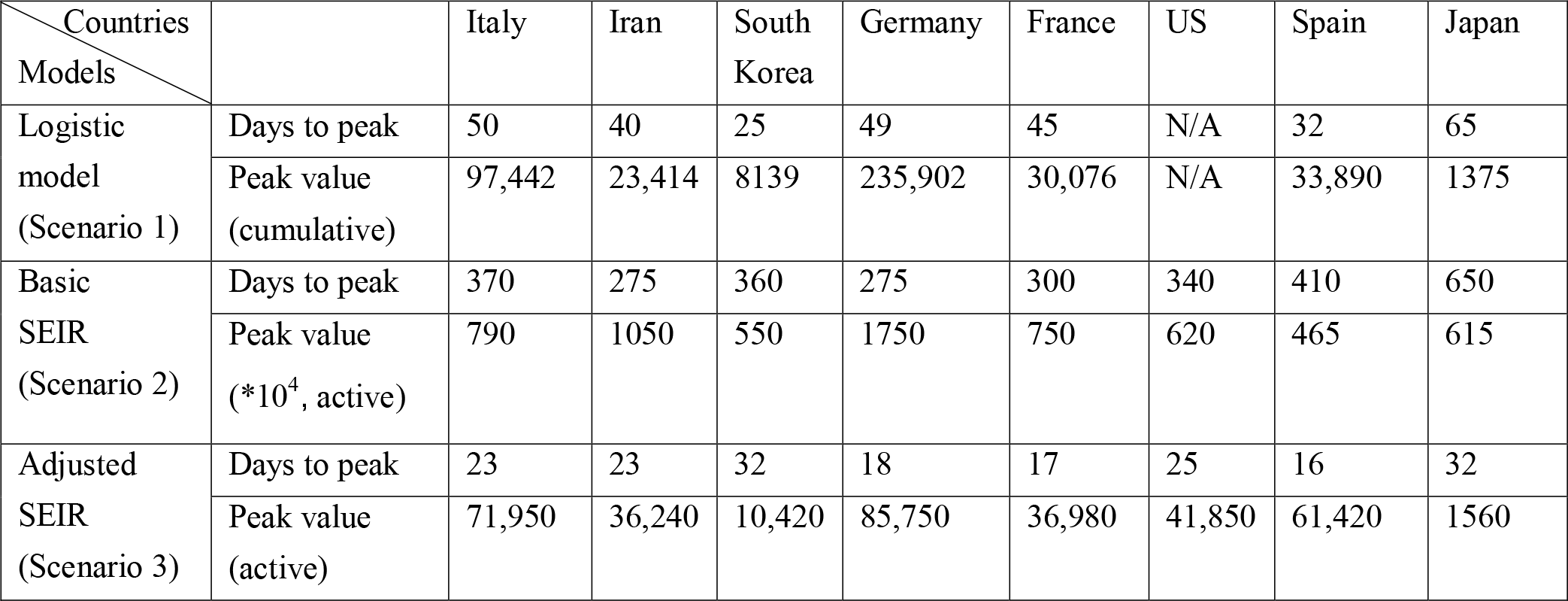
Epidemic peak predictions of the 8 countries

| Countries<br>Models |  | Italy | Iran | South<br>Korea | Germany | France | US | Spain | Japan |
| --- | --- | --- | --- | --- | --- | --- | --- | --- | --- |
| Logistic<br>model<br>(Scenario 1) | Days to peak | 50 | 40 | 25 | 49 | 45 | N/A | 32 | 65 |
|  | Peak value<br>(cumulative) | 97,442 | 23,414 | 8139 | 235,902 | 30,076 | N/A | 33,890 | 1375 |
| Basic<br>SEIR<br>(Scenario 2) | Days to peak | 370 | 275 | 360 | 275 | 300 | 340 | 410 | 650 |
|  | Peak value<br>(*10<sup>4</sup>, active) | 790 | 1050 | 550 | 1750 | 750 | 620 | 465 | 615 |
| Adjusted<br>SEIR<br>(Scenario 3) | Days to peak | 23 | 23 | 32 | 18 | 17 | 25 | 16 | 32 |
|  | Peak value<br>(active) | 71,950 | 36,240 | 10,420 | 85,750 | 36,980 | 41,850 | 61,420 | 1560 |

In Scenario 1, based on the logistic model, we predicted the epidemic trends of the 8 countries. Table 1 and Appendix B list the detailed results of the prediction and growth trajectories of the 8 countries. For instance, the model placed the peak time of Italy as 50 days after its initial date of February 23, 2020, with a maximum number of infected individuals of approximately 97,442. The infection of US was unable to be predicted based on a logistic model using reported data because it is exponentially growing, which makes it difficult to fit the logistic curve, which reflects that the US is in a stage of high spread of the disease.

In Scenario 2, the basic SEIR results showed that the confirmed cases will take 9-22 months (275-650 days) to reach their peak, and most of the population would eventually be infected over a long period of time if there are no control measures. Appendix C lists the detailed results of the basic SEIR model predictions of different countries. In addition, the active number of cases at the peak time will reach approximately 10-20% of these countries’ populations, thus overloading the healthcare system, which is the worst possible scenario, as shown in Table 2.

In Scenario 3, the adjusted SEIR model results show that under strict control measures, the number of active cases will reach a peak in 16-32 days (from early April to middle April 2020) after the initial date, when the number of cases reaches 100. Appendix D shows the detailed results of the adjusted SEIR model predictions of the 8 countries. Japan and South Korea took isolation measures when the disease was still in the early stage of transmission, its peak value was low, and the cumulative number of infected people was relatively small. Especially in Japan, our predicted peak value is less than 1560, which shows that the disease spread has been well controlled in the early stage. However, the transmission in European countries and the US is in the outbreak phase as of the date of our data collection. The adjusted SEIR model predicted that the peak values of Spain, Italy, Germany, France and other countries are between 10,420 and 85,750.

According to our parameter estimation methods, we disclosed the dynamics of the number of cases. The Scenario 3 estimation is based on the strict quarantine assumption, and the results showed that implementing the control measures would decrease the epidemic peak significantly and bring it forward. This supports the conclusion that the contact rate is an important factor that reflects the effects of control measures; with the formulation and implementation of extreme epidemic prevention measures to reduce the rate of contact, the epidemic size and peak would decrease. However, the epidemic still shows a long tail after the peak, and our study found that a longer quarantine period of susceptibility would reduce the long tail after the peak.

### Spreading Potentials

The growth rates of the logistic curves are also listed as a spreading potential index for comparing the situations of the 8 countries in Table 3. Our results suggest that these countries all have a high risk of rapid virus transmission except for Japan, where the spread is slowing down. On the basis of evidence from previous transmission dynamics studies, the documented COVID-19 basic reproductive number (R_0_) ranges from 2.0 to 4.9^17-20^. Our estimated R_0_ range is between 1.687∼3.864 (with the assumption of a 7-day mean infection period in terms of reported COVID-19 studies).

**Table 3.**
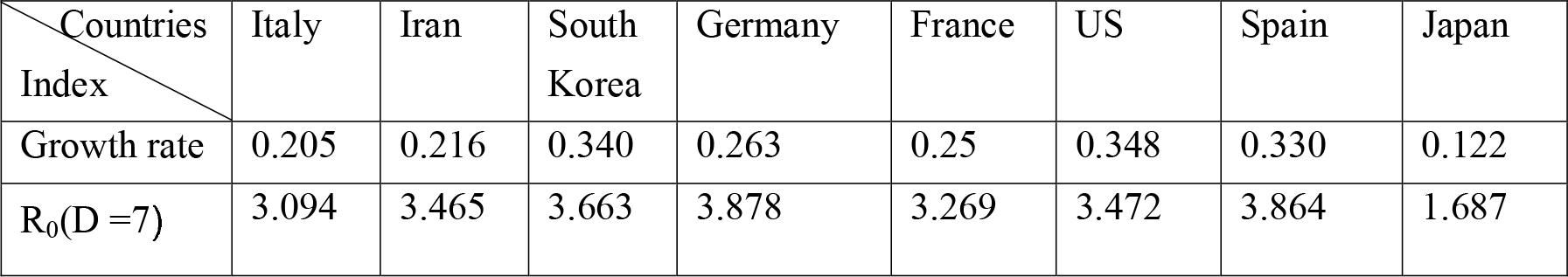
Spreading potential of the 8 countries

| Countries<br>Index | Italy | Iran | South<br>Korea | Germany | France | US | Spain | Japan |
| --- | --- | --- | --- | --- | --- | --- | --- | --- |
| Growth rate | 0.205 | 0.216 | 0.340 | 0.263 | 0.25 | 0.348 | 0.330 | 0.122 |
| $R_0(D=7)$ | 3.094 | 3.465 | 3.663 | 3.878 | 3.269 | 3.472 | 3.864 | 1.687 |

## Discussion

According to the COVID-19 data tracking, these 8 countries have delayed the phase of preventing the epidemic and have indeed entered the outbreak phase with community-spread cases. Mortality rate analysis disclosed that undetected transmission events may have occurred in some countries. Seven of the eight countries have R0 levels above 3, which deserves our attention.

The high mortality was reported because the main reason for undetected transmission is probably that some of the cases are asymptomatic or lack testing kits. Our Scenario 2 and Scenario 3 are separately based on maximal assumption (without any interventions) and minimal assumption (with strict interventions). In Scenario 3, under the assumption that infected people are promptly tested and counted, the adjusted model suggested that the peak time will be reached after the strict implementation of two large-scale rounds of 14 days of quarantine. The decrease in the epidemic size in Japan and South Korea showed positive evidence, as shown in Table 1. However, considering that each country has different cultures and healthcare situations and that the implementation of the policies and control measures are at different levels^21^, the actual situation of those countries should be between Scenario 2 and Scenario 3. It is worth pointing out that, compared with implementing no interventions, if governments take strict control measures to reduce the movement of the population and implement prompt diagnosis and isolation, the peak time will be reached, and the peak size would greatly decrease to a relatively low level in approximately 30 days; it is best triggered in the early stage of the epidemic; however, it will need to be maintained for several months until a vaccine becomes available. Conversely, as long as the epidemic continues, there is still a possibility of future outbreaks if governments loosen interventions and allow people to return to close social distances.

The results of spreading potentials show that the R_0_ of COVID-19 is high, which is similar to that of SARS viruses (R_0_ values between 2.0 and 5.0^22^), higher influenza viruses H1N1 (R_0_ values between 1.2 and 3.7^23^), and Ebola viruses (R_0_ values between 1.34 and 3.65^24^). COVID-19 is a highly contagious human-to-human transmission disease. The R_0_ is expected to decrease substantially compared to values at the early stage after governments implement control measures; however, regardless of the kind of policy each country executes, each policy has its limitations in defending against COVID-19, and sustained transmission chains will occur until there is a vaccine or the until the virus disappears due to seasonal or population immunity^25^. Therefore, detecting all transmission events is the most critical issue of COVID-19 control in the current stage because any undetected case in a local area could begin a new epidemic chain of transmission. In addition, the public should take adequate protective measures against the transmission of COVID-19.

From the view of mathematical models, the SEIR model is designed for infectious disease estimation; however, the logistic growth model is designed to fit the development of the curves. The logistic model may fit the existing data better when compared with the SEIR model, since it is trained from the existing data, but it cannot be accurately judged and incorporates infectious characteristics. Therefore, we believe that the logistic model is better for near-term predictions. On the other hand, the SEIR model introduces more variables and factors by considering the interaction and association among multiple groups of people, and it is more reasonable than the logistic model as it follows the rules of infectious disease development, but the prediction results vary greatly with respect to different interventions and settings.

The study has some limitations. The mathematical models allow for the quick incorporation of multiple inputs to yield prediction results. However, this process involves making assumptions about uncertain factors; for example, it is difficult to determine the exact extent to which people follow the local government’s quarantine policies or measures and engage in behaviors such as washing hands, using masks, and social distancing. This may affect the actual contact rate and the subsequent development of the epidemic. The models also lack enough data to estimate the quarantine proportions of a certain population. In fact, the evolution of the epidemic is quite complicated, and our study has only taken into account a few factors. In addition, a lack of testing kits means many cases have not been tested in some countries, and without robust testing, the official number of cases is incomplete. When working with incomplete data, a small error in one factor can have an outsize effect.

In conclusion, our study demonstrated that reducing the contact rate is a key measure in controlling the spread of disease at an early stage, and spending enough time in quarantine would decrease the scale of cases after the peak. Therefore, implementing strong containment policies during the early spreading stages of COVID-19 and flattening the peak to avoid overloading the healthcare system should be listed as the main actions in these high-risk countries/regions. After the strict quarantine period, governments still need to raise public awareness of precautions and the importance of engaging in self-protective behaviors to bring the epidemic under control as a series of scattered events.

## Data Availability

Contact Prof. Yun Long or Prof. Shuyang Zhang to access the data.

## References

1 Organization, W. H. Director-General’s remarks at the media briefing on 2019-nCoV on 11 February 2020, <https://www.who.int/dg/speeches/detail/who-director-general-s-remarks-at-the-media-briefing-on-2019-ncov-on-11-february-2020 (> (2020).

2 Prevention, C. o. D. C. a. Coronavirus Disease 2019 (COVID-19), < https://www.cdc.gov/coronavirus/2019-ncov/prepare/transmission.html?CDC_AA_refVal=https%3A%2F%2Fwww.cdc.gov%2Fcoronavirus%2F2019-ncov%2Fabout%2Ftransmission.html > (

3 WHO Director-General’s opening remarks at the media briefing on COVID-19 - 11 March 2020, <https://www.who.int/dg/speeches/detail/who-director-general-s-opening-remarks-at-the-media-briefing-on-covid-19---11-march-2020> (2020).

4 Hermanowicz, S. W. Forecasting the Wuhan coronavirus (2019-nCoV) epidemics using a simple (simplistic) model. medRxiv, doi:10.1101/2020.02.04.20020461. (2020).

5 Liu, T. et al. Transmission dynamics of 2019 novel coronavirus (2019-nCoV). (2020).

6 Imai, N. et al. Report 2: Estimating the potential total number of novel Coronavirus cases in Wuhan City, China. Imperial College London (2020).

7 Yang, Z. et al. Modified SEIR and AI prediction of the epidemics trend of COVID-19 in China under public health interventions. Journal of Thoracic Disease (2020).

8 Hong, N. et al. Evaluating the secondary transmission pattern and epidemic prediction of the COVID-19 in metropolitan areas of China. medRxiv (2020).

9 Dong, E., Du, H. & Gardner, L. An interactive web-based dashboard to track COVID-19 in real time. Lancet Infect Dis, doi:10.1016/S1473-3099(20)30120-1 (2020).

10 Wu, K., Darcet, D., Wang, Q. & Sornette, D. Generalized logistic growth modeling of the COVID-19 outbreak in 29 provinces in China and in the rest of the world. arXiv preprint 2003.05681 (2020).

11 Pell, B., Kuang, Y., Viboud, C. & Chowell, G. Using phenomenological models for forecasting the 2015 Ebola challenge. Epidemics 22, 62–70 (2018).

12 Tian, H. et al. An investigation of transmission control measures during the first 50 days of the COVID-19 epidemic in China. Science, eabb6105, doi:10.1126/science.abb6105 (2020).

13 Countries in the world by population (2020), <https://www.worldometers.info/world-population/population-by-country/> (2020).

14 Organization, W. H. Coronavirus disease 2019 (COVID-19) Situation Report, <https://www.who.int/docs/default-source/coronaviruse/situation-reports/20200219-sitrep-30-covid-19.pdf> (2020).

15 Van den Driessche, P. & Watmough, J. in Mathematical epidemiology 159–178 (Springer, 2008).

16 Diekmann, O., Heesterbeek, J. & Roberts, M. G. The construction of next-generation matrices for compartmental epidemic models. Journal of the Royal Society Interface 7, 873–885 (2010).

17 Zhao, S. et al. Preliminary estimation of the basic reproduction number of novel coronavirus (2019-nCoV) in China, from 2019 to 2020: A data-driven analysis in the early phase of the outbreak. International Journal of Infectious Diseases (2020).

18 Wu, J. T., Leung, K. & Leung, G. M. Nowcasting and forecasting the potential domestic and international spread of the 2019-nCoV outbreak originating in Wuhan, China: a modelling study. Lancet, doi:10.1016/S0140-6736(20)30260-9. (2020).

19 Shen, M., Peng, Z., Xiao, Y. & Zhang, L. Modelling the epidemic trend of the 2019 novel coronavirus outbreak in China. bioRxiv (2020).

20 Park, S. W. et al. Reconciling early-outbreak estimates of the basic reproductive number and its uncertainty: framework and applications to the novel coronavirus (SARS-CoV-2) outbreak. medRxiv (2020).

21 Yang, X. et al. Transportation, Germs, Culture: A Dynamic Graph Model of 2019-nCoV Spread. (2020).

22 Liu, Y., Gayle, A. A., Wilder-Smith, A. & Rocklöv, J. The reproductive number of COVID-19 is higher compared to SARS coronavirus. Journal of travel medicine (2020).

23 Boni, M. F. et al. Modelling the progression of pandemic influenza A (H1N1) in Vietnam and the opportunities for reassortment with other influenza viruses. BMC Med 7, 43, doi:10.1186/1741-7015-7-43 (2009).

24 House, T. Epidemiological dynamics of Ebola outbreaks. Elife 3, e03908 (2014).

25 Ferguson, N. M. et al. Impact of non-pharmaceutical interventions (NPIs) to reduce COVID-19 mortality and healthcare demand. London: Imperial College COVID-19 Response Team, March 16 (2020).

